# Pragmatic trial design of a digital supportive care platform for patients with brain tumours and their carers

**DOI:** 10.64898/2026.08.18.26360754

**Authors:** Sarah CE Bray, Verena Schadewaldt, Meinir Krishnasamy, James R Whittle, Wendy Chapman, Kit Huckvale, Kara Burns, Daniel Capurro, Meredith J Layton, Joseph Thomas, Richard De Abreu Lourenco, Diana Andrew, Heidi McAlpine, Rana S Dhillon, Sarah Cain, Mark Rosenthal, Katharine J Drummond, Mahima Kalla

## Abstract

Patients with a brain tumour receive evidence-based clinical care in Australia but a focus on supportive care, including social connection, is often deficient. Digital health platforms hold promise to support these patients and their carers. Existing platforms often lack end-user co-design, evidence-based development and rigorous evaluation. Recognising this unmet need, we co-designed Brain Tumours Online, a digital supportive care platform to streamline access to educational resources, symptom management tools, and peer support for patients, carers, and healthcare professionals. In this article, we present our evaluation approach for Brain Tumours Online to advance methodological thinking in the evaluation of multi-faceted, co-designed digital health platforms. In contrast to standardised procedures in clinical trials, digital health interventions such as supportive care platforms are more complex due to their interactive nature, no prescriptive protocols for usage and the dynamic content of web-based information. Thus, traditional evaluation approaches often fall short in evaluating such multi-faceted digital health supportive care platforms. To address these challenges, we developed a bespoke, logic-modelling based evaluation approach to assess the usability, engagement, impact, and economic value of our platform. Our pragmatic but rigourous evaluation approach required the adaptation of existing evaluation frameworks, subject-matter, and lived experience expert knowledge. Our implementation science and co-design approach are shared in different papers. Our study outcomes will also be shared in a separate paper. In the current paper, we share our approach to the evaluation of Brain Tumours Online and provide insights that may be of value for other researchers interested in the nuances of trialing multi-faceted digital health supportive care platforms.

## Introduction

Brain tumours are a heterogenous group of neoplasms with variable survival depending on age and tumour type. Regardless of the tumour type and aggressiveness, patients with a brain tumour and their carers experience long-term impacts on their physical, psychosocial, cognitive and financial quality of life^1–3^. These challenges are compounded by isolation, which can be a result of disability, geographic distance from specialist centres^4,5^ and the rarity of brain tumours. Supportive care needs are insufficiently addressed by existing models of care. Digital health platforms may help fill this gap in unmet supportive care needs, especially for patients who face barriers to accessing services, such as geographical location. Digital health platforms can range from the provision of digital information via websites or apps, to electronic devices such as pedometers, with the goal to improve health outcomes ^6^.

### The Brain Tumours Online platform

Previously, there was no comprehensive, Australian evidence-informed digital supportive care platform for patients with brain tumours and their carers. Available resources tended to feature ad-hoc development and implementation with limited patient engagement, digital health expertise, or underpinning evidence base^7–10^. Based on a decade of research documenting the quality of life of Australian patients with brain tumours^1,11^, and establishing the need for digital support in disease management for patients with a brain tumour^8,12^, the authors co-designed Brain Tumours Online (https://braintumoursonline.org) ^13–15^.

Brain Tumours Online is a digital supportive care platform to streamline access to evidence-based educational resources, digital health tools and connection for patients affected by a brain tumour and their carers. The platform features three pillars - Learn, Connect, and Toolbox (Figure 1), each of which addresses an unmet need. The Learn pillar provides comprehensive, evidence-based information on brain tumours, their effects, treatments, and support services. Trustworthy resources are delivered within a curated, searchable directory which can be filtered by tumour type, stage of illness, geographical location, and user type (patient, carer, or healthcare professional). The Connect pillar allows patients and carers to engage with each other and healthcare professionals in a number of ways including sharing personal stories, connecting with others through a social media platform via the Mighty Networks App, and participating in live or recorded interactive webinars with subject-matter experts and people with lived experience. The Toolbox pillar features validated digital symptom-management therapeutic tools, including an evidence-based cognitive behavioural therapy program to improve sleep (Sleep Healthy Using The Internet; SHUTi)^16^, and an Australia-based online carer support intervention (CarerWell)^17^.

**Figure 1.**
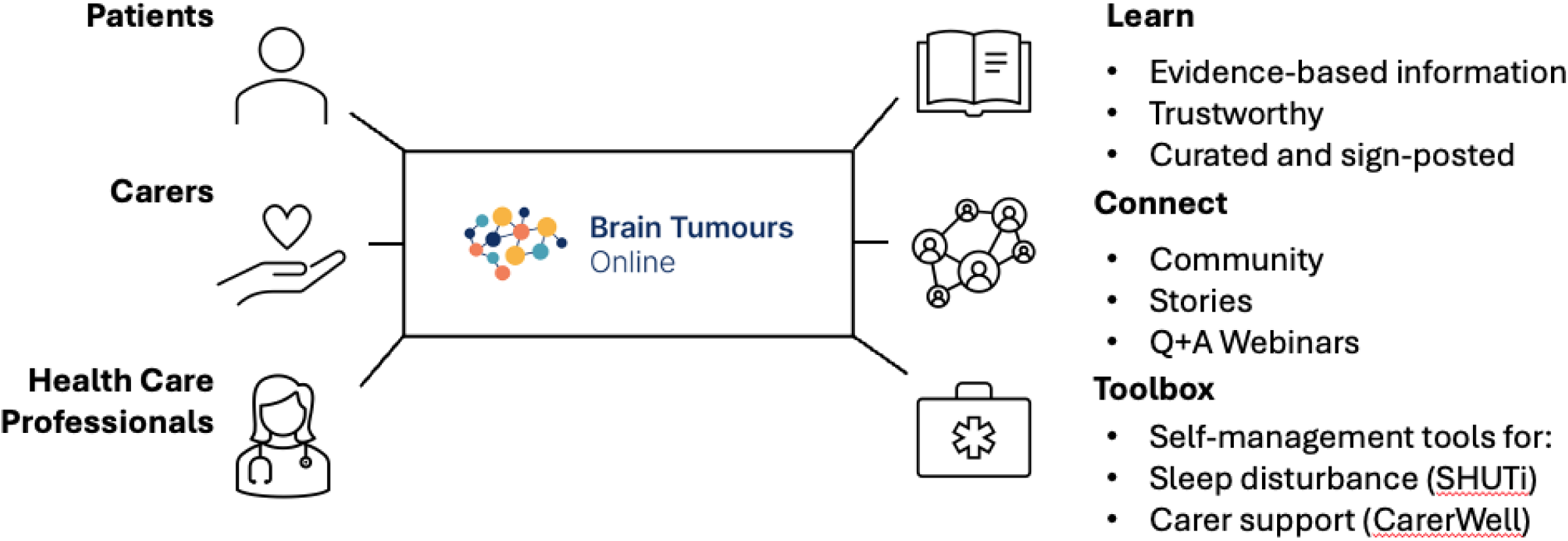
The three user groups and the main components of Brain Tumours Online.

Once co-designed and developed, our intention was to rigorously evaluate Brain Tumours Online, to generate evidence about its usability, usefulness and impact for different users (patients, carers and health care professionals), and to develop recommendations to iteratively improve the platform in response to end-user feedback. In developing our evaluation approach for Brain Tumours Online, we came across a variety of challenges.

### Evaluating digital health interventions

The World Health Organization (WHO) provides guidance on monitoring and evaluation of digital health interventions^18^. WHO suggests a range of evaluation activities depending on the stage of evaluation (e.g. usability, efficacy, and effectiveness). However, this guidance is largely indicative, rather than prescriptive, resorting mostly to a menu of methodological options from which to choose. In the absence of a gold standard methodological approach, we set out to develop a bespoke evaluation program for Brain Tumours Online. To this end, we conducted a series of activities to synthesise knowledge from research literature, lived experience expertise of patients and carers, and clinicians’ domain knowledge to determine what implementation success for Brain Tumours Online will entail.

One key challenge our team faced was to develop a fit-for-purpose, pragmatic, but academically rigorous evaluation approach, which would be capable of generating actionable insights to support the long-term translation of this platform in the real-world. In this paper, we present our evaluation approach for Brain Tumours Online with the aim of advancing the methodological discourse on trialling of digital health platforms to support real-world translational success. In addition, we have previously also published two other related methodological papers as part of this project, a) our implementation perspective and strategy, including the evidence-base informing it ^19^; and b) our stakeholder engagement and co-design approach^14^.

### Our evaluation planning process

We undertook a series of activities towards developing our evaluation plan, as illustrated in Figure 2. The steps conducted included: 1) Develop logic model; 2) Conduct literature review; 3) Determine success criteria; 4) Prioritise criteria for evaluation; and 5) Finalise the evaluation plan.

**Figure 2:**
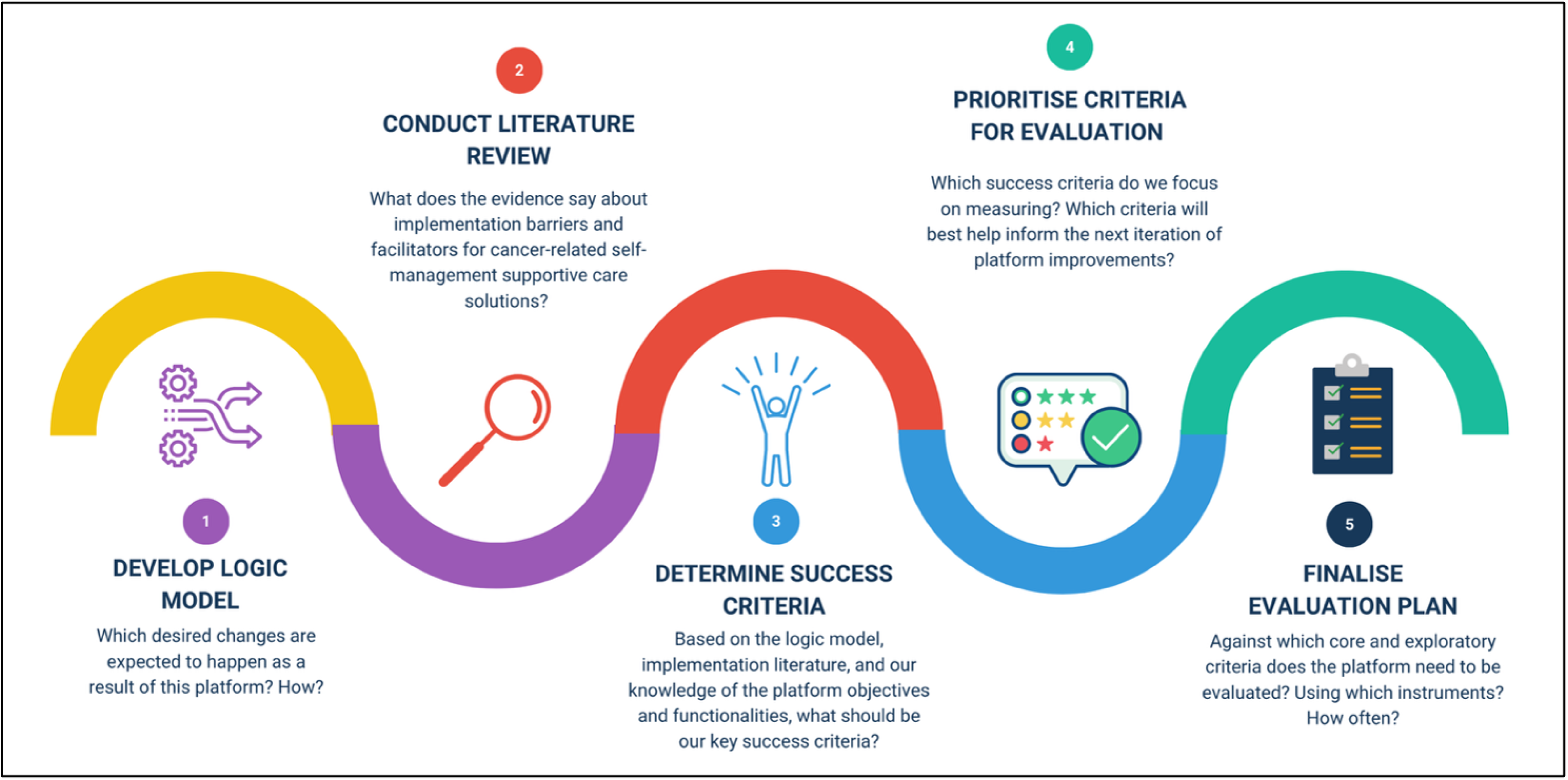
Evaluation Planning Approach.

### Step 1 – Develop logic model

Logic models are a useful means of describing how a program or complex intervention works. Logic models can be helpful for developing a shared understanding about a program’s inputs, activities, planned outputs, and anticipated outcomes^20,21^. They can also help surface assumptions regarding the underlying mechanisms of change within a project or complex intervention, and are increasingly being utilised for quality improvement and implementation science^20^.

Given the multifaceted nature of the Brain Tumours Online platform, we commenced our logic modelling exercise, with three simple questions:

1. Is the platform usable? *(‘Usability’ - If the platform is not usable, people will be unlikely to use it)*
2. Do people use the platform? *(‘Engagement’ – If the platform is usable, do people engage with it and use it?)*
3. Do users benefit from the platform? *(‘Impact’ – If the platform is usable, and people engage with it, do they benefit from it?)*

Based on these three overarching themes of ‘usability’, ‘engagement’, and ‘impact’, a logic model was co-produced by the project stakeholders. First, a small working group of digital health researchers (author names redacted for peer review) came together to develop a draft logic model for the platform. Subsequently, the draft logic model was shared with the broader team as part of an evaluation planning workshop. During this workshop, other project stakeholders, including clinicians, researchers and lived experience experts gave their inputs and helped finesse the logic model (Figure 3).

**Figure 3:**
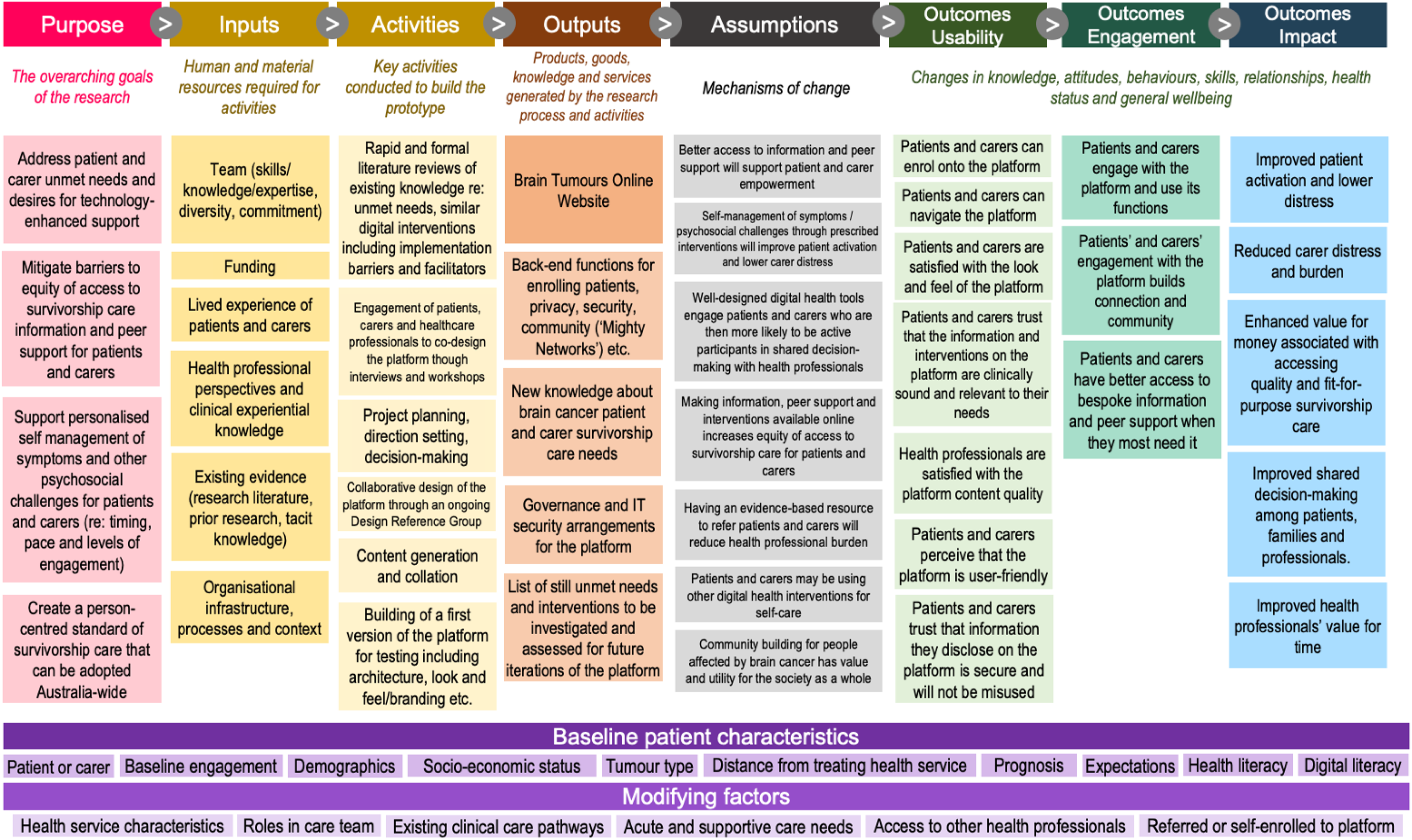
Brain Tumours Online Platform Logic Model.

### Step 2 – Conduct literature review

Greenhalgh and Russell note that evaluations of digital health initiatives often fail to yield meaningful insights^22^. Given the socio-technical contextual complexities of implementing and trialing digital health innovations, traditional assumptions, methods, and study designs associated with the experimental science paradigm fall short within digital health evaluation^22^. Thus, we sought to take an implementation science led approach when developing the evaluation plan for Brain Tumours Online. To this end, we embarked on a literature review which identified 10 themes relating to implementation barriers and facilitators for other cancer self-management digital health platforms. The insights drawn from this literature review built the bases for the development of our bespoke implementation strategy^15^ and were synthesized into success criteria for Brain Tumours Online, as discussed in the following section.

### Step 3 – Determine success criteria

Based on the platform logic model developed in *Step 1* and implementation literature insights derived from *Step 2*, author (name redacted for blind peer review) prepared an initial suite of potential success criteria for the Brain Tumours Online platform. The findings from Steps 1 and 2 were distilled into 30 success criteria across seven overarching dimensions: 1) Platform quality, innovation, usability; 2) Accessibility; 3) Holistic and personalised features; 4) Awareness of platform features and belief in benefit; 5) Data security, legal, liability; 6) Healthcare professional interface / involvement; 7) User training and ability to use the platform effectively.

### Step 4 - Prioritise criteria for evaluation

The next step involved prioritisation of the success criteria towards identifying key outcomes of interest for the platform’s evaluation. We conducted an online prioritisation workshop with project stakeholders including clinicians, digital health researchers, health economists, industry development partner, patient and carer representatives. The workshop was facilitated by author (name redacted for blind peer review) who had prior experience conducting co-production and stakeholder engagement activities. First, all workshop attendees individually categorised the overarching seven evaluation dimensions identified in step 3, as ‘Essential’, ‘Nice to have’, or ‘Not required’. Subsequently, the attendees were divided into breakout groups, with each group assigned 2-3 evaluation dimensions. Each group was asked to review the various success criteria within their assigned dimensions and allocate each criterion to ‘Essential’, ‘Nice to have’, or ‘Not required’ categories. A summary of key stakeholder inputs received during the workshop has been provided in Supplementary Material 5. This process resulted in 21 final shortlisted success criteria for the platform, based on what stakeholders considered Essential (Table 1).

**Table 1:**
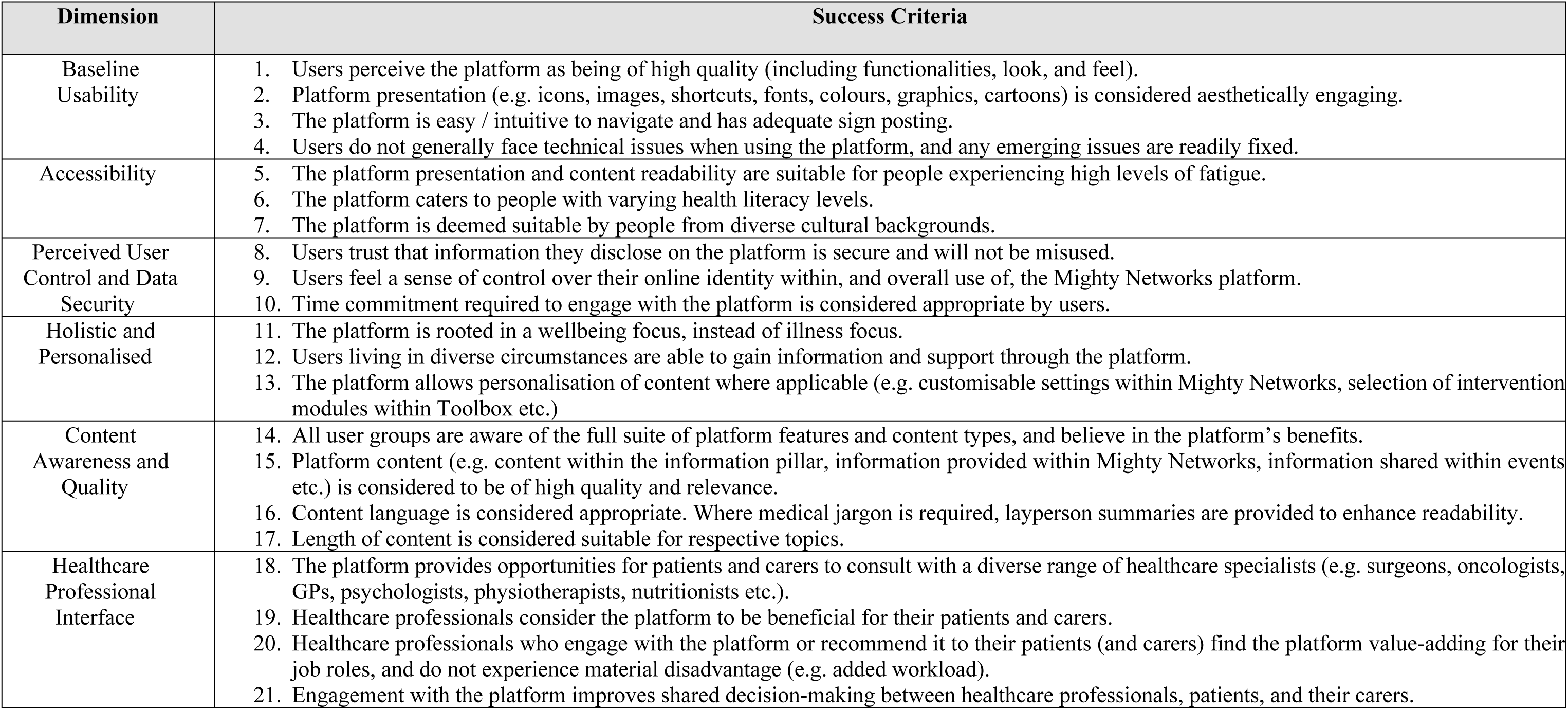
Platform Success Criteria for Brain Tumours Online.

### Step 5 – Finalisation of evaluation plan

Having reached consensus on what implementation success for the Brain Tumours Online platform would entail, we then proceeded to establish our evaluation research questions and corresponding data collection and analysis methods in the form of a study protocol.

### Evaluation Study Characteristics

#### Design and methodology

Overall, our evaluation program was conceptualised as a concurrent mixed-methods, single-arm study, with a separate health economic analysis component. The study design was based on a pragmatist epistemological positioning, which steers clear of metaphysical debates about the nature of reality and instead focuses on the generation of actionable findings^23^. Given the real-world nature of our study, this epistemological stance was considered suitable. Pragmatist research often includes mixed methods study designs with multiple methods being considered complementary to each other for generation of meaningful, actionable insights^23^.

As per the platform logic model described previously, we had identified three outcome categories: usability, engagement, and impact. Thus, the evaluation plan’s research questions and corresponding data collection methods were established according to these three outcome categories. In addition, specific evaluation aims and data collection methods were articulated for the SHUTi and CarerWell digital therapeutic tools included within the *Toolbox* pillar of the Brain Tumours Online platform. Lastly, a health economic value assessment study was developed as part of the overarching evaluation plan. Chosen data collection methods included: 1) an initial enrolment survey to capture demographic information, prognosis, treatments and expectations of the platform, etc.; 2) a range of validated Participant-reported Outcome Measures (PROMs - usually ‘P’ refers to “Patients” but we are using “Participant” in this study as we intended to collect these measures from both patients and carers) with impact on unmet needs (CASUN/CASPUN tools) being the primary outcome; 3) a final exit survey based on the platform success criteria described previously; 4) semi-structured exit interviews; 5) web analytics usage data for the Brain Tumours Online platform using Google analytics and web scraping tools; 6) event-specific feedback surveys for webinars hosted within the *Connect* pillar of the platform; and 7) de-identified Pharmaceutical Benefits Scheme and Medicare data for health economics analysis. A summary of the various components of the evaluation program, research questions, and corresponding data collection method has been provided in Table 2.

**Table 2:**
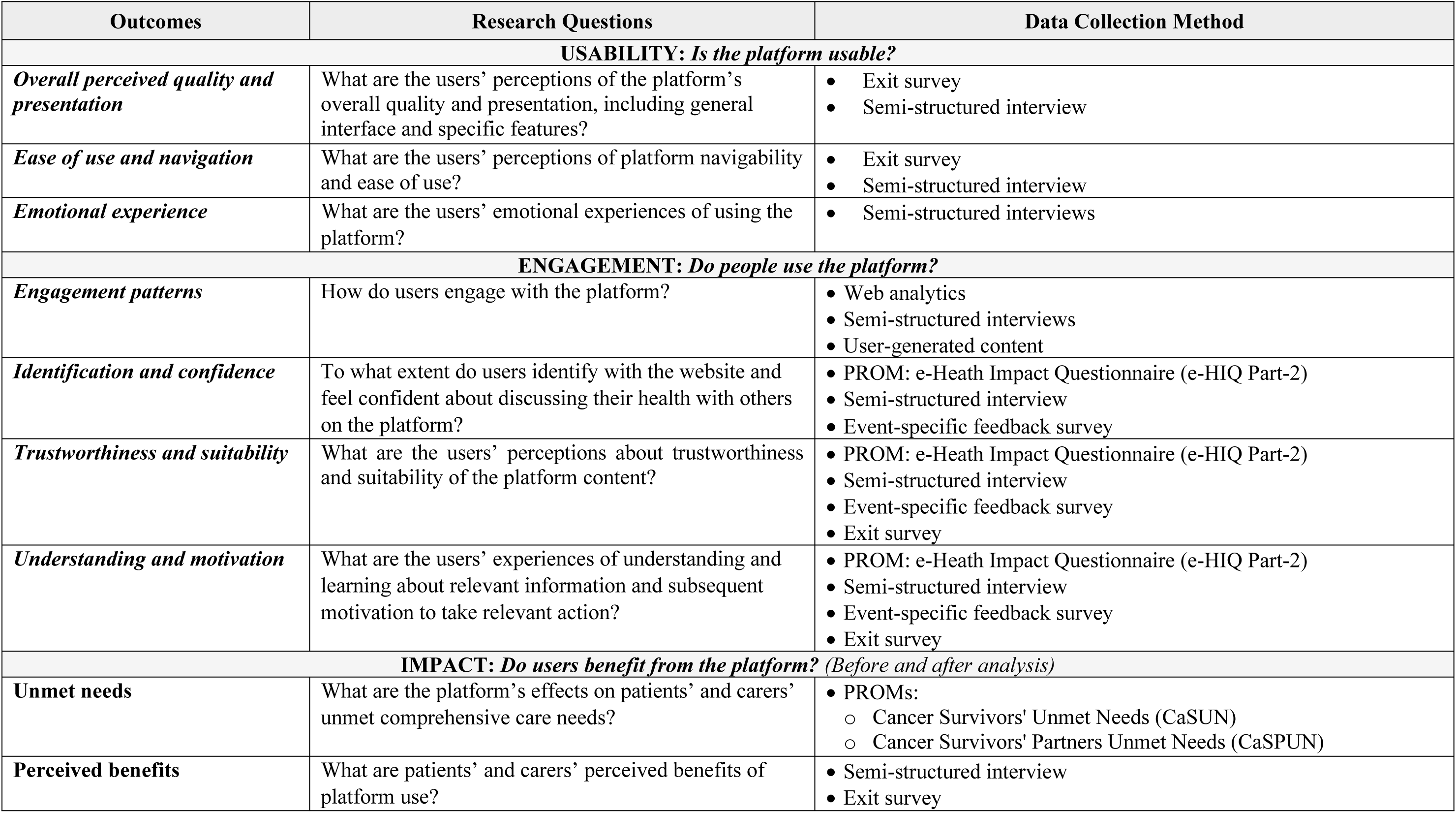
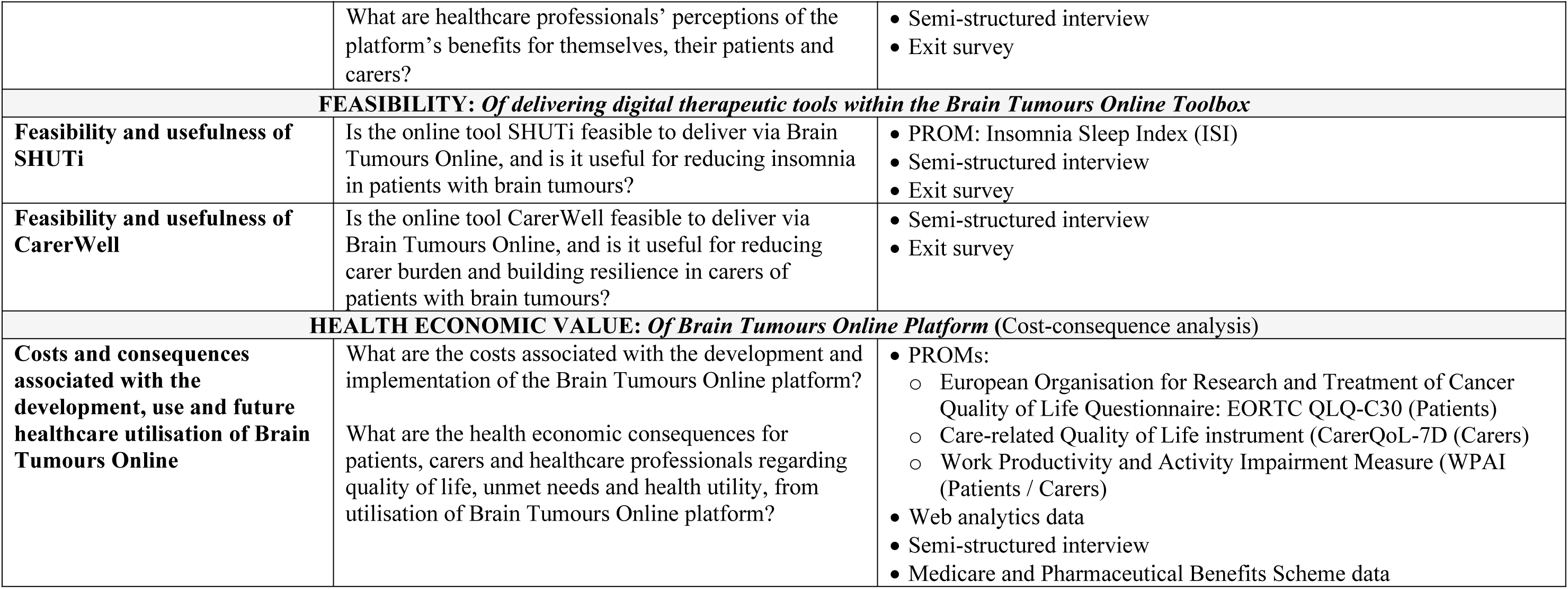
Summary of study outcomes, research questions, and data collection methods.

### Study population and selection criteria

We planned to recruit three participant groups to evaluate Brain Tumours Online:

- **Patients**: adults with a primary brain tumour;
- **Carers**: current or bereaved family members/friends acting as informal (unpaid) carers for an adult with a primary brain tumour; and
- **Health Care Professionals**: those whose work includes care of adults with primary brain tumours and/or their carers.

All participants needed to be aged 18 years and older, live in Australia, able to communicate in English and have access to an internet-enabled electronic device. Patients with secondary brain tumours (metastases) and formal (paid) carers were not eligible to participate.

### Recruitment and sampling

Recruitment would be conducted over a 6-month period. Participants were to be recruited either: 1) by the clinician investigators at one of four participating hospitals during a clinical consultation or via email; 2) via social media, professional networks, advocacy groups, presentations at conferences, clinical seminars or consumer engagement events; or 3) via referral from other participants or self-referral from the Brain Tumours Online website landing page.

To join Brain Tumours Online, applicant study participants had to verify their eligibility and complete online informed consent. Eligible participants who provided consent, would then receive baseline surveys (Supplementary Material 1) via REDCap for online completion. Upon completion of the baseline surveys, participants would receive instructions to create a login and password to access Brain Tumours Online. Usage data would be collected over the evaluation period, and additional surveys and requests for feedback (for example via interview) sent out to participants at various timepoints throughout the study. At the end of the evaluation period, access would remain open to participants beyond completion of the formal data collection window, in anticipation of further modifications as a result of the evaluation, and transition to a publicly accessible version of the platform.

The World Health Organization’s model for the evaluation of digital health interventions recommends recruitment of at least 100 participants to ensure robust evaluation of feasibility, usability and impact^18^. Our multi-modal recruitment strategy was designed to recruit a diverse cross-section of participants in Australia with varied sociodemographic characteristics, tumour types, and experiences, ensuring a wide breadth of perspectives.

### Data collection

Quantitative and qualitative data were planned to be collected at baseline, 3- and 6-month intervals, and at activity-specific time points. Data collection methods included user surveys and interviews; PROMs to identify changes over time; data on health service use to inform the health economic analysis; and component-specific evaluations of resources delivered through the Toolbox and Connect pillars. Web analytic data on participant interactions with the platform were to be collected continuously from first login to the end of study period to gather quantitative user metrics. Passive collection of web analytics data minimised the burden on participants and decreased reliance on memory to self-report usage. Survey and PROMs data would be captured in REDCap directly from participants. Semi-structured interviews (Supplementary Material 3) would be conducted towards the end of the evaluation period with a diverse sub-sample across the three participant groups by phone or video conference and recorded for transcription and analysis.

## Ethics declaration

The study was approved by the Royal Melbourne Hospital Human Research Ethics Committee on 22/12/2022 (HREC/89522/MH-2022). This ethics committee is constituted and operated in accordance with the National Statement on Ethical Conduct in Human Research 2023. A summary of all data collection methods, relevant participant groups, and timepoints is provided in Table 3.

**Table 3:** Data collection methods, timepoints, and relevant participant groups. *HCP = Health Care Professional

| Data collection method | Patient | Carer | HCP* | Baseline | 3-months | 6-months | Activity specific |
| --- | --- | --- | --- | --- | --- | --- | --- |
| **Enrolment survey** to collect demographic data and expectations of the platform (Supplementary Material 1) | x | x | x | x |  |  |  |
| **Exit survey** to collect end-of-study feedback on overall experience and perceived benefits of the platform (Supplementary Material 2) | x | x | x |  |  | x |  |
| **Web analytics** to identify patterns of user engagement and attrition | x | x | x | Continuously collected |  |  |  |
| **PROM: e-Health Impact Questionnaire (e-HIQ, Part 1)** to assess general attitudes towards using the internet to access health information<sup>41</sup> | x | x |  | x |  |  |  |
| **PROM: e-Health Impact Questionnaire (e-HIQ, Part 2)** to assess ease with using online information<sup>41</sup> | x | x |  |  |  | x |  |
| **PROM: European Organisation for Research and Treatment of Cancer (EORTC) QLQ-C30** to assess quality of life specific to a cancer population<sup>42</sup> | x |  |  | x | x | x |  |
| **PROM: Cancer Survivors' Unmet Needs (CaSUN)** to assess unmet needs in patients impacted by a brain tumour<sup>43</sup> | x |  |  | x | x | x |  |
| **PROM: Work Productivity and Activity Impairment (WPAI)** to assess the impact of sickness on the capacity to conduct usual activities, including work<sup>44</sup> | x | x |  | x | x | x |  |
| **PROM: Cancer Survivors' Partners Unmet Needs (CaSPUN)** to assess unmet needs in partners who care for a person with a brain tumour<sup>45</sup> |  | x |  | x | x | x |  |
| **PROM: Care-related Quality of Life instrument (CarerQoL-7D)** to assess the impact of caregiving on general well-being and quality of life<sup>46</sup> |  | x |  | x | x | x |  |
| **Qualitative semi-structured interviews:** to collect in-depth feedback about users' experiences with Brain Tumours Online, including engagement with the social media network, and the toolbox (Supplementary Material 3) | x | x | x |  |  | x |  |

## Data analysis

Analyses were planned to assess the usability, engagement, impact, feasibility and health economic value of the platform. In line with our mixed methods study design, data from some sources would be analysed individually with results then compared across multiple sources within our study^24^. Thus, findings in one area could provide important explanatory context for the interpretation of data elsewhere, for example if qualitative interview statements support or contradict quantitative usage from web analytics or PROMs data^25^.

In other instances, data from disparate sources would be combined in one database and analysed together to provide a more complete picture of the usefulness and impact of the platform. Engagement data from web analytics and the social media platform Mighty Networks would be extracted and entered into the same database to allow a more comprehensive understanding of how participants used the platform and its third-party features as a whole.

### Analysis of enrolment and exit surveys

Descriptive statistics would be reported to summarise enrolment and exit survey results (Supplementary material 1 and 2). Enrolment surveys would collect participant socio-demographic and disease characteristics, including age, gender, language spoken at home, postcode of residence, treating service, tumour type (for patients), relationship to patient (for carers) and specialty (for HCP). In the exit surveys participants would be asked to rate aspects of their user experience including technical issues, data control and security, content, presentation, liked and disliked features. Likert-type scale ratings on platform quality and usability would be presented using median and range. Proportions and percentages would be used to report categorical data. Feedback in exit surveys would be cross-checked with interview reports to verify and corroborate findings.

### Qualitative analysis of interviews

Initial interviews would be analysed while further interviews were being conducted until thematic saturation is reached. This usually occurs after 9-17 interviews in homogenous groups^26^. Accounting for heterogeneity in our sample, we therefore aimed to interview up to 40 participants (i.e., 15 patients, 15 carers and 10 HCP) to allow for differing perspectives and ensure thematic saturation within each group.

The transcription software Otter.ai^27^ would be used to generate verbatim interview transcripts, followed by a quality check and de-identification by a research team member. Transcripts would be managed and thematically analysed with Nvivo^28^ using a combination of inductive and deductive approaches^29^. An inductive approach provides a nuanced picture on participants’ perception of the platform. The deductive approach is based on specific success criteria developed for this project and their reflection in participant interviews^29^. Two members of the project team would independently code a subset of interview transcripts for each participant group (patients, carers, health care professionals), then compare codes to ensure rigour. Disagreements would be resolved through consultation with a third researcher. Data triangulation by contrasting exit survey feedback and web-analytics with subjective user reports in interviews would be applied to better interpret findings.

### Analysis of Web Analytics Data

Statistical analysis of web analytics data - including the number of user sessions, frequency and duration of engagement with particular content, common search terms and filters, engagement with the online community, and attrition – would be conducted to summarise patterns of user engagement with Brain Tumours Online. Interview data would be used to corroborate emergent patterns. Correlations between user engagement with the platform and their PROMs would be estimated using multiple linear regression.

### Analysis of PROMs

The primary impact assessments (CASUN, CASPUN) would allow identification of any change in participants’ unmet needs over the duration of the study. PROMs would be assessed based on the difference between outcomes at baseline and the three- and six-month timepoints. Items would be scored according to each instrument’s respective guidelines, with descriptive statistics used to summarise and evaluate the potential impact of any missing data.

Analyses would focus on investigating the relationships between users’ engagement with the platform and their PROMs, while controlling for participants’ relevant background characteristics. Linear mixed models would be used to estimate correlations, with PROMS treated as dependent variables and platform engagement and participant socio-demographic characteristics treated as independent (control) variables. Fixed effects would be applied for time and platform engagement; random effects applied for participant outcomes. Data would also be stratified for sub-group analysis based on emergent patterns and data availability.

### Analysis of engagement patterns

Latent profile analysis, a statistical method, would be conducted to identify user groups with similar engagement patterns based on their shared characteristics such as socio-demographic details, unmet needs, activity impairment, quality of life and overall level of engagement.

### Analysis of User-Generated Content

Within the ‘Connect’ pillar of the platform, users generate their own content, including social media posts and responses; writing, sharing and commenting on stories and submitting questions and suggesting topics for webinar events. To evaluate how users engage in the social media community, data scraping would be employed to identify frequency of post, comments and views.

### Analysis of Tool-Specific Data

Qualitative feedback about user experience for the tools (SHUTi and CarerWell) would be analysed thematically as part of interviews (and, where appropriate, exit surveys). Web analytics would also be used to present descriptive statistics on tool-specific user patterns and completion rates.

For SHUTi, Insomnia Severity Index (ISI)^30^ scores was used to determine changes related to using the tool. Previous studies have used as few as 10 participants to establish feasibility and preliminary efficacy^31^. Clinically meaningful improvement has been determined as a reduction of at least six points on the ISI^32^. Statistical significance of pre- and post-SHUTi scores would be possible by a paired t-test analysis.

### Analysis of Event-Specific Feedback

Event-specific feedback surveys (Additional file 4) were designed to capture perceptions of the online webinars, and suggestions for future topics. Quantitative data from rating scales were summarised using descriptive statistics and presented as frequencies. Qualitative data from free text fields were summarised thematically and compared (and integrated with interview or exit survey data when appropriate). Review of data from feedback surveys throughout the evaluation period would also be used to inform the development of subsequent webinars based on feedback from the participants.

### Health Economic Analysis

The health economic value of Brain Tumours Online would be assessed through a ‘cost-consequence analysis’, a form of health economic analysis describing the costs and health outcomes of available treatment alternatives^33^. Consequences would be analysed as the extent to which engagement with the platform is associated with: 1) a change in public health care service use and 2) participants’ engagement in usual activities, relative to the pre-intervention period. Changes in public health service use would be assessed using de-identified patient-level data from the Medical Benefits Schedule (MBS) and Pharmaceutical Benefits Scheme (PBS), two Australian Government programs that subsidise medical services and medicines. Changes in participation in usual activities among patients and carers will be assessed using the Work Productivity and Activity Impairment (WPAI) tool. Assessment of costs will focus on the costs to develop and implement the platform such as infrastructure requirements and time (including value of time spent by HCP to deliver content through the platform); ‘time-toxicity’ (i.e., the value of time spent by participants to engage with the platform); and the costs of public health care service use among patients and carers. Health service costs would be assessed as the value of public health benefits (i.e., MBS and PBS) paid, plus out-of-pocket costs to participants (the difference between the published cost of services and government benefits paid). Quality of life questionnaires completed by patients (EORTC-QLQ C30) and carers (Carer QoL) would also be expressed as health utilities for inclusion in future economic evaluations^34,35^.

## Discussion

We outlined the methodological process of generating a bespoke evaluation plan for our digital supportive care platform Brain Tumours Online. The strength of our approach is the theory-led, stakeholder-informed and literature-based development of the evaluation plan on which the pragmatic steps for study design, data collection and analysis are based. Key features of our design are collaboration with patients and carers, using subjective/qualitative feedback from users in combination with objective/quantitative data to evaluate usability, engagement, impact, feasibility and health economic value of Brain Tumours Online. The results from this evaluation will not only provide a thorough evaluation of the usability, engagement and impact of this digital health platform, but will inform iterative and future content, technical and design improvements. This flexibility of continuous improvement, even during the study period, added to the complexity of evaluating Brain Tumours Online and we summarise our reflections on the challenges of evaluating digital health platforms below.

### Challenges of evaluating Brain Tumours Online

Digital health platforms hold tremendous potential to improve health and wellbeing of patients. Historically, randomised controlled trial designs have been considered the gold standard for evaluating the effectiveness of pharmacological and other health interventions. These designs are well suited when an intervention and its effects can be clearly causally linked and measured^36^. In contrast, digital health platforms such as Brain Tumours Online have significantly greater complexity due to their interactive nature, including on-demand use as per personal need and preference, no specific ‘prescription’ for use of platform, and the dynamic nature of the content^37^. Unlike clinical treatment trials, where participants follow standardised protocols, participants’ use of the platform during the study period was self-directed, mimicking a real-world situation. Thus, data evaluation is challenging, with less standardised participant data for benchmarking and comparison.

When developing the evaluation approach for Brain Tumours Online, our team was cognizant that participants would be able to engage with the platform in a variety of ways to meet their individual needs, including being able to choose which aspects of the platform they wanted to use and what frequency and intensity, based on their own preferences, health and supportive care needs. We anticipated that usage type and frequency may vary between patients, carers and health care professionals. Additionally, among patients and carers, usage may vary based on the patients’ tumour types and disease stages (for example early diagnosis, during treatment, advanced stages). Similarly, due to the online and self-guided nature of the study, participants unable to navigate or use some features, or with low digital literacy, may be less likely to provide feedback during the evaluation period, reducing the opportunity for researchers to identify barriers for this population.

Another challenge was that the platform would not remain static during the evaluation period. As per the original intent of our co-design activities, the platform was designed with the view to constantly evolve in response to user feedback. Based on participants’ engagement in the evaluation phase, the platform would evolve in response to users’ feedback and requests for content via direct and indirect input garnered through community posts and participant contributions. In particular, features such as resources contained within the ‘Learn’ pillar, social media content on the Might Networks online peer community, and webinars would continue to expand even as the platform was being evaluated.

The complexities of evaluating the platform were further compounded by considerations associated with brain tumours, wherein survivorship of some tumour types is low, illness prognosis poor, and trajectories complex. All of these factors would influence participant attrition. Additionally, given the fortunately lower number of diagnosed brain tumour cases compared to other cancers, reaching participant numbers to achieve statistical power in a randomised study was thought to be challenging. Furthermore, participants had to complete a series of online PROMs and surveys at baseline before being provided access to the platform, and questionnaire completion is a known burden in human research^38,39^. This may have posed an artificial barrier to participation and use of the Brain Tumours Online platform during the evaluation phase, which would not be the case in a real-world non-research setting. Completion of a series of online surveys may be a particular challenge for people with a lower digital literacy, or experiencing fatigue or decreased cognitive abilities. We recognise this as a potential recruitment and retention challenge.

We also anticipated challenges when offering resources delivered through third party software/platforms. Unlike secure dedicated research tools such as REDCap, commercial third-party tools, such as Zoom for hosting webinars and Mighty Networks for the social media community, do not provide researchers with full control of participant data submission, collection, storage and use. Managing the use of third-party programs is challenging from a research governance perspective, both to mitigate and reduce risks, but also to inform participants of that which is outside of the researchers’ control. This may have limited some participant engagement, which we will investigate during the evaluation.

On the other hand, we required our evaluation approach to generate actionable insights about potential usability issues, illuminate patterns of user engagement, identify what the different user groups - healthcare professionals, patients, carers - desired of the platform, and how they each benefited from it. Additionally, the health economic value proposition of the platform also needed to be ascertained, especially given its objective to bridge currently unmet supportive care needs. Thus, the evaluation would need to generate salient insights to help inform future decisions around platform expansion, care model and funding decisions^40^.

## Conclusion

Reflecting on the evaluation plan process and documenting the evaluation of a digital health platform adds to the existing literature and provides information for the development and evaluation of future digital health platforms. With the increase in digital health platforms available it is crucial to ascertain a thorough evaluation prior to releasing it to users. Too many health apps and online platforms have not been tested or created with consumer input. Brain Tumours Online is the first Australian digital health platform and resource for patients with a brain tumour and their families, that has been developed with extensive consumer engagement, digital health expertise, HCP input and is evidence-based. The platform is now publicly available (https://braintumoursonline.org) with future modifications to be incorporated based on evaluation findings.

## Data Availability

No datasets were generated or analysed during the current study. All relevant data from this study will be made available upon study completion.

## Acknowledgements

We thank the patients, carers, consumer advocates, health care professionals and researchers who have contributed to this research and the development of Brain Tumours Online through their input in meetings, workshops and interviews. We appreciate their valuable time spent on this study. The project described in this paper is supported by the Digital Health Validitron, a collaborative and interdisciplinary research group that assists digital health innovators from healthcare, academia and industry to accelerate the creation of evidence that proves the real-world value of their ideas and products. This study received funding from the Australian Government through the Medical Research Future Fund 2020 Brain Cancer Survivorship Grant Program (2021-2024) (ID: MRFBII000014). The funder had no role in the execution of the study.

## Author Contributions

SCEB: Conceptualization, Data curation, Formal analysis, Investigation, Methodology, Project administration, Resources, Writing – original draft, Writing – review & editing VS: Conceptualization, Data curation, Formal analysis, Investigation, Methodology, Resources, Writing – original draft, Writing – review & editing

MKy: Conceptualization, Funding acquisition, Investigation, Methodology, Supervision, Writing – review & editing

JRW: Conceptualization, Funding acquisition, Investigation, Methodology, Writing – review & editing

WC: Conceptualization, Investigation, Methodology, Supervision, Writing – review & editing

KH: Conceptualization, Investigation, Methodology, Supervision, Writing – review & editing

KB: Conceptualization, Investigation, Methodology, Writing – review & editing

DC: Conceptualization, Investigation, Methodology, Writing – review & editing

MJL: Conceptualization, Investigation, Methodology, Project administration, Writing – review & editing

JT: Conceptualization, Investigation, Methodology, Writing – review & editing

JDAL: Conceptualization, Funding acquisition, Investigation, Methodology, Supervision, Writing – review & editing,

DA: Conceptualization, Investigation

HM: Conceptualization, Investigation, Methodology, Writing – review & editing

RSD: Conceptualization, Investigation, Methodology, Writing – review & editing

SC: Conceptualization, Investigation, Methodology, Writing – review & editing

MR: Conceptualization, Funding acquisition, Investigation, Methodology, Writing – review & editing

KJD: Conceptualization, Funding acquisition, Investigation, Methodology, Supervision, Resources, Writing – review & editing

MK: Conceptualization, Data curation, Formal analysis, Investigation, Methodology, Project administration, Resources, Validation, Writing – original draft, Writing – review & editing

## References

1 Teng, K. X. et al. Life after surgical resection of a low-grade glioma: A prospective cross-sectional study evaluating health-related quality of life. J Clin Neurosci 88, 259–267 (2021). 10.1016/j.jocn.2021.03.038

2 Haider, S., Taphoorn, M. J. B., Drummond, K. J. & Walbert, T. Health-related quality of life in meningioma. Neurooncol Adv 3, vdab089-vdab089 (2021). 10.1093/noajnl/vdab089

3 Tallant, J., Pakzad-Shahabi, L., Lambert, S. D., Williams, M. & Wells, M. Quality of Life in Caregivers of Patients with Brain Tumours: A Systematic Review and Thematic Analysis. European Journal of Cancer Care 2023, 2882837 (2023). 10.1155/2023/2882837

4 Mallya, S. et al. A qualitative analysis of the benefits and barriers of support groups for patients with brain tumours and their caregivers. Support Care Cancer 28, 2659–2667 (2020). 10.1007/s00520-019-05069-5

5 Janda, M. et al. Unmet supportive care needs and interest in services among patients with a brain tumour and their carers. Patient Educ Couns 71, 251–258 (2008). 10.1016/j.pec.2008.01.020

6 Kampmeijer, R., Pavlova, M., Tambor, M., Golinowska, S. & Groot, W. The use of e-health and m-health tools in health promotion and primary prevention among older adults: a systematic literature review. BMC Health Serv Res 16 **Suppl 5**, 290 (2016). 10.1186/s12913-016-1522-3

7 Fridriksdottir, N., Gunnarsdottir, S., Zoëga, S., Ingadottir, B. & Hafsteinsdottir, E. J. G. Effects of web-based interventions on cancer patients’ symptoms: review of randomized trials. Support Care Cancer 26, 337–351 (2018). 10.1007/s00520-017-3882-6

8 McAlpine, H., Joubert, L., Martin-Sanchez, F., Merolli, M. & Drummond, K. J. A systematic review of types and efficacy of online interventions for cancer patients. Patient Educ Couns 98, 283–295 (2015). 10.1016/j.pec.2014.11.002

9 Schaefer, I. et al. Quality of online self-management resources for adults living with primary brain cancer, and their carers: a systematic environmental scan. BMC Palliative Care 20, 22 (2021). 10.1186/s12904-021-00715-4

10 Zheng, C. et al. Benefits of Mobile Apps for Cancer Pain Management: Systematic Review. JMIR Mhealth Uhealth 8, e17055 (2020). 10.2196/17055

11 Nassiri, F. et al. Life after surgical resection of a meningioma: a prospective cross-sectional study evaluating health-related quality of life. Neuro Oncol 21, i32–i43 (2019). 10.1093/neuonc/noy152

12 McAlpine, H., Sejka, M. & Drummond, K. J. Brain tumour patients’ use of social media for disease management: current practices and implications for the future. Patient Education and Counseling 104, 395–402 (2021).

13 Kalla, M. et al. Co-Designing a User-Centered Digital Health Tool for Supportive Care Needs of Patients With Brain Tumors and Their Caregivers: Interview Analysis. JMIR cancer 11, e53690 (2025).

14 Kalla, M. et al. To framework, or not to framework? Reflections from co-design of a digital supportive care platform for patients with brain tumours and their carers. Digital Health 11, 20552076251339302 (2025).

15 Schadewaldt, V. et al. Development of an evidence-informed implementation strategy for a digital supportive care platform for brain tumour patients, their carers and healthcare professionals. Digital Health 11, 20552076251316713 (2025).

16 Ritterband, L. M. et al. Effect of a Web-Based Cognitive Behavior Therapy for Insomnia Intervention With 1-Year Follow-up: A Randomized Clinical Trial. JAMA Psychiatry 74, 68–75 (2017). 10.1001/jamapsychiatry.2016.3249

17. Kay-Lambkin, F. CarerWell, <https://carerwell.com.au/> (2024).

18 WHO. 133 (World Health Organization, Geneva, 2016).

19 Schadewaldt, V. et al. Development of an evidence-informed implementation strategy for a digital supportive care platform for brain tumour patients, their carers and healthcare professionals. Digital Health 11, 20552076251316713 (2025).

20 Seidl, K. L. et al. Development of a logic model to guide implementation and evaluation of a mobile integrated health transitional care program. Population health management 24, 275–281 (2021).

21 Savaya, R. & Waysman, M. The logic model: A tool for incorporating theory in development and evaluation of programs. Administration in Social Work 29, 85–103 (2005).

22 Greenhalgh, T. & Russell, J. Why do evaluations of eHealth programs fail? An alternative set of guiding principles. PLoS medicine 7, e1000360 (2010).

23 Morgan, D. L. Pragmatism as a paradigm for social research. Qualitative inquiry 20, 1045–1053 (2014).

24 Moran-Ellis, J. et al. Triangulation and integration: processes, claims and implications. Qualitative Research 6, 45–59 (2006). 10.1177/1468794106058870

25 Fetters, M. D., Curry, L. A. & Creswell, J. W. Achieving integration in mixed methods designs - principles and practices. Health Services Research 48, 2134–2156 (2013). 10.1111/1475-6773.12117

26 Hennink, M. & Kaiser, B. N. Sample sizes for saturation in qualitative research: A systematic review of empirical tests. Social science & medicine 292, 114523 (2022).

27. Otter.ai, <https://otter.ai/> (2024).

28. Lumivero. NVivo (Version 14). (2022).

29 Braun, V. & Clarke, V. Using thematic analysis in psychology. Qualitative research in psychology 3, 77–101 (2006).

30 Morin, C. M., Belleville, G., Bélanger, L. & Ivers, H. The Insomnia Severity Index: psychometric indicators to detect insomnia cases and evaluate treatment response. Sleep 34, 601–608 (2011). 10.1093/sleep/34.5.601

31 Mattos, M. K. et al. Feasibility and Preliminary Efficacy of an Internet-Delivered Intervention for Insomnia in Individuals with Mild Cognitive Impairment. J Alzheimers Dis 84, 1539–1550 (2021). 10.3233/JAD-210657

32 Yang, M., Morin, C. M., Schaefer, K. & Wallenstein, G. V. Interpreting score differences in the Insomnia Severity Index: using health-related outcomes to define the minimally important difference. Curr Med Res Opin 25, 2487–2494 (2009). 10.1185/03007990903167415

33. Michael F. Drummond, Mark J. Sculpher, George W. Torrance, Bernie J. O’Brien & Stoddart, G. L. Methods for the economic evaluation of health care programme. 3rd edn, (Oxford University Press, 2005).

34 Hoefman, R. J., van Exel, J. & Brouwer, W. B. F. Measuring Care-Related Quality of Life of Caregivers for Use in Economic Evaluations: CarerQol Tariffs for Australia, Germany, Sweden, UK, and US. Pharmacoeconomics 35, 469–478 (2017). 10.1007/s40273-016-0477-x

35 King, M. T. et al. Australian Utility Weights for the EORTC QLU-C10D, a Multi-Attribute Utility Instrument Derived from the Cancer-Specific Quality of Life Questionnaire, EORTC QLQ-C30. Pharmacoeconomics **36**, 225-238 (2018). 10.1007/s40273-017-0582-5

36 Blandford, A. et al. Seven lessons for interdisciplinary research on interactive digital health interventions. Digital health 4, 2055207618770325 (2018).

37 Karpathakis, K. et al. An evaluation service for digital public health interventions: user-centered design approach. Journal of medical Internet research 23, e28356 (2021).

38 Kunz, T. & Gummer, T. Effects of objective and perceived burden on response quality in web surveys. International Journal of Social Research Methodology: Theory & Practice, No Pagination Specified-No Pagination Specified (2024). 10.1080/13645579.2024.2393795

39. Yan, T., Fricker, S. & Tsai, S. in Advances in Questionnaire Design, Development, Evaluation, and Testing (eds Paul C. Beatty et al.) Ch. 8, 193-212 (John Wiley & Sons Inc, 2020).

40 De Santis, K. K., Jahnel, T., Mergenthal, L., Zeeb, H. & Matthias, K. Evaluation of digital interventions for physical activity promotion: protocol for a scoping review. JMIR Research Protocols 11, e35332 (2022).

41 Kelly, L., Jenkinson, C. & Ziebland, S. Measuring the effects of online health information for patients: Item generation for an e-health impact questionnaire. Patient Education and Counseling 93, 433–438 (2013). 10.1016/j.pec.2013.03.012

42 Aaronson, N. K. et al. The European Organization for Research and Treatment of Cancer QLQ- C30: a quality-of-life instrument for use in international clinical trials in oncology. J Natl Cancer Inst 85, 365–376 (1993). 10.1093/jnci/85.5.365

43 Hodgkinson, K. et al. The development and evaluation of a measure to assess cancer survivors’ unmet supportive care needs: the CaSUN (Cancer Survivors’ Unmet Needs measure). Psychooncology 16, 796–804 (2007). 10.1002/pon.1137

44 Reilly, M. C., Zbrozek, A. S. & Dukes, E. M. The validity and reproducibility of a work productivity and activity impairment instrument. Pharmacoeconomics 4, 353–365 (1993). 10.2165/00019053-199304050-00006

45 Hodgkinson, K. et al. Assessing unmet supportive care needs in partners of cancer survivors: the development and evaluation of the Cancer Survivors’ Partners Unmet Needs measure (CaSPUN). Psycho-Oncology 16, 805–813 (2007). 10.1002/pon.1138

46 Brouwer, W. B., van Exel, N. J., van Gorp, B. & Redekop, W. K. The CarerQol instrument: a new instrument to measure care-related quality of life of informal caregivers for use in economic evaluations. Qual Life Res 15, 1005–1021 (2006). 10.1007/s11136-005-5994-6

